# Clinical and genetic predictors of dementia in Parkinson’s disease

**DOI:** 10.64898/2026.03.06.26347693

**Authors:** Matthew R Solomons, Naomi Hannaway, Oliver Fox, Alyssa Costantini, Raquel Real, Angelika Zarkali, Huw R. Morris, Rimona S Weil, the Vision in Parkinson’s Study team

## Abstract

**Importance:** Dementia is common in Parkinson’s disease (PD), causing greater disability than other symptoms, but varies in timing. Although visual deficits are linked with PD dementia, how these interact with genetic factors to predict PD dementia has not been characterised.

**Objective:** To investigate whether visual deficits and genetic factors predict PD dementia.

**Design:** Large prospective longitudinal case-control study, mean follow-up 32.7 (SD=12.3) months.

**Setting:** Cases were recruited between 2017-2020 at 35 UK PD clinics.

**Participants:** People with PD without dementia at baseline were included.

**Main outcomes and measures:** Visual function was measured using a web-based platform. The main outcome measure was global cognition, measured as the Montreal Cognitive Assessment (MoCA). Blood samples were collected for genetics.

**Results:** 450 patients with PD were included. Mean age of PD patients was 71.7 (SD=7.8), 68% male. Mean baseline MoCA was 27.7 (SD=1.7). 263 patients with PD were classed as poor-vision based on baseline visual tests: mean age 74.4 (SD=6.8) compared to 69.7 (SD=7.5) with good-vision. Poor-vision PD patients had higher rates of progression to mild cognitive impairment (PD-MCI) (HR=2.34, CI=1.58-3.48, *p*FDR=0.00062, age- and sex-corrected). The combination of genetic factors together with vision influenced outcomes. In good-vision PD patients, high-risk *GBA1* gene variants were linked with greater progression to PD-MCI (HR=4.61, CI=1.73-12.28, *p*FDR=0.0068). Polygenic Risk Score (PRS) for both PD and Alzheimer’s disease (AD) also modified cognitive survival when combined with vision status. High PD-PRS was associated with greater progression to PD-MCI in good-vision patients (HR=2.66, CI=1.21-5.81, *p*FDR=0.0381); and high AD-PRS with greater progression to PD-MCI in poor-vision PD patients (HR=1.91, CI=1.10-3.32, *p*FDR=0.04999). Combining high PD- and AD-PRS, compared to low PD- and AD-PRS in good-vision PD showed even higher progression to PD-MCI (HR=6.14, CI=1.36-27.83, *p*FDR=0.046). Simulations showed that adding visual and genetic stratification reduced sample size from n=705 to n=160 for clinical trials.

**Conclusions and relevance:** Poor vision in PD predicts progression to PD-MCI and dementia. This combines with the effects of genetic factors including *GBA* risk variants and PD- and AD-PRS. These findings can enable enrichment of clinical trials for patients at higher risk of PD dementia, for more efficient trial design for interventions to slow progression.

**Key points:** *Question:* Do clinical factors, measured by performance on visual tests, and genetic factors help predict which patients are more likely to develop cognitive involvement in Parkinson’s disease?

*Findings:* This prospective longitudinal study of 450 Parkinson’s patients, based in Parkinson’s clinics, with mean follow-up 32.7 months, found that Parkinson’s patients with poor vision are more likely to develop cognitive impairment; and that genetic factors in combination with poor vision further predict poor prognostic groups for Parkinson’s dementia.

*Meaning:* These data could enable selection of Parkinson’s patients at highest risk of dementia for clinical trials aimed at slowing Parkinson’s dementia.

## Introduction

Parkinson’s disease (PD) is the fastest growing neurological condition worldwide, with 7 million people currently affected, growing to an estimated 14 million by 2040^1^. Although conceptualised as a movement disorder, dementia is common in PD, with cognitive deficits affecting nearly half of people with PD within 10 years’ of diagnosis^2^. When cognition impacts on day-to-day activities in PD, it is known as PD dementia (PDD) and is characterised by deficits in visuo-perceptual, executive and attention domains^3^. PDD is associated with a more aggressive disease trajectory and increased risk of mortality^4,5^. It represents an important milestone in disease progression, impacts on quality-of-life, and is a key event in clinical trials for PD. PD with mild cognitive impairment (PD-MCI) is where cognitive impairment is present, but does not interfere with day-to-day activities. It usually predates PDD.

We currently lack disease-modifying treatments for PD and PDD. However, new approaches providing biological classifications for PD are opening new opportunities for clinical trials^6,7^. There is increased understanding of the importance of co-pathologies in PDD, with beta-amyloid and tau strongly implicated in driving disease progression^8,9^. With approvals for anti-amyloid therapies for Alzheimer’s disease (AD) there is more focus on considering these therapies for co-pathologies in PDD^10^, but a limitation for clinical trials is the need to include large numbers.

A key challenge in PD is to predict which patients are more likely to develop dementia. Early identification will enable them to be targeted for disease modifying treatments, as these emerge. Enriching for fast progressors to PD dementia will improve trial efficiency, allowing smaller numbers to be included^11^. Whilst age is the main risk factor for dementia in PD, there remains variability in timing of PDD onset, even when adjusting for age^12^. Other factors linked with rapid progression to dementia include male sex, REM sleep behaviour disorder, depression, autonomic dysfunction, and abnormalities on DAT-SPECT scans and CSF abeta and tau^13,14^.

There is now converging evidence from several centres that the presence of visual dysfunction in PD is linked with higher risk of developing dementia and poor outcomes: Patients with PD who make errors copying intersecting pentagons have nearly double the risk of dementia by 10-year follow-up^2^; patients with impaired colour vision are more likely to develop dementia^15^; and people with PD with poorer vision are more likely to develop dementia, frailty or death at three-year follow-up^16^. At population level, people with PD who also have visual deficits are more likely to develop poor outcomes including dementia, over time^17^. However, so far this has only been examined prospectively in smaller studies, and we lack large-scale prospective data that poorer visual function predicts cognitive decline in PD.

Several genetic risk factors have also been identified as contributing to risk of dementia in PD. These include variants in the *GBA1* gene^18^, rare monogenic mutations in *SNCA*^19^, and mixed findings on the contribution of the *APOE4* allele^20–23^. Additional genetic risk loci for progression to dementia were identified using two recent time-to-event genome-wide association studies (GWAS)^24,25^. Information from GWAS risk variants can be combined using polygenic risk scores (PRS), calculated using a weighted sum of risk alleles from each carrier, based on the effect size in the GWAS^26^. Previous work has suggested a link between case-control PD-PRS and cognitive decline in PD^27^.

How the presence of visual deficits in PD combines with genetics to predict risk of progression to dementia is not yet known. Smaller-scale studies with detailed measures of visual dysfunction are not powered to detect genetic differences between groups; and large-scale genetic studies have not tested visual function.

The goal of the current study was therefore to establish whether visual deficits predict cognitive change over time in PD, how these combine with genetic factors; and whether combining this information could enable smaller sample sizes to be included in clinical trials to slow progression of cognitive decline in PD. We examined visual function in a large prospective real-world study of nearly 500 people with PD attending PD clinics. We used a web-based platform validated in PD to test visual function, collected blood samples for genetic testing, and examined cognition at yearly intervals, up to 4-year follow-up.

## Methods

### Participants and procedures

The Queen Square Research Ethics Committee approved the study. Informed consent was obtained from all participants according to the Declaration of Helsinki. We followed EQUATOR reporting guidelines.

Participants were recruited between March 2016 and March 2020 from 35 participating PD clinics in the UK. Inclusion criteria were clinically-diagnosed PD according to Movement Disorders Society guidelines^28^ (early- to mid-stage disease, Hoehn & Yahr 1-3), within 10 years’ diagnosis. Exclusion criteria included atypical Parkinsonism, age-of-onset less than forty-five years, age at start of study older than 81, cognitive involvement at baseline visit (defined as MoCA less than 25 at baseline), patient-reported ophthalmological disease, psychiatric disease (except anxiety or depression) and confounding neurological conditions. Participants attended at baseline, and then annually for three years. See Figure 1 for flow chart of participants included in the study. At each session, participants underwent assessment of global cognition and visuo-perceptual assessments using a web-platform.

**Figure 1.**
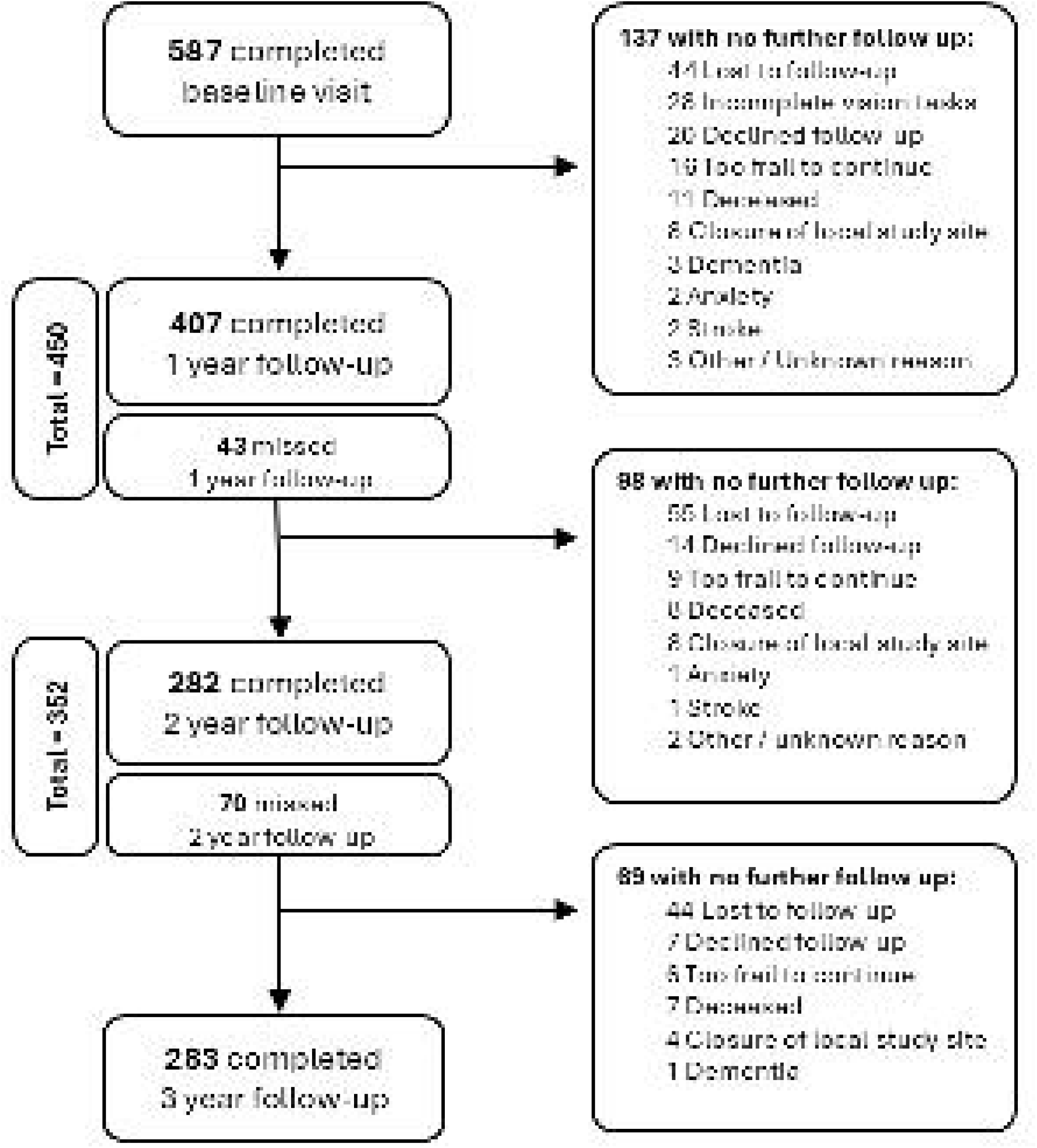
Flow chart showing patients recruited and tested in the study at baseline, and during follow-up. Reasons for exclusion and loss to follow-up are given at each stage. Where patients are lost to follow-up due to PD-dementia, these are included in outcomes. PD, Parkinson’s disease

Global cognition was assessed using the MoCA version 7.1.^29^ For participants unable to attend in person for follow-up visits during the COVID-19 pandemic, the MoCA was conducted through video calling (with video-share for full MoCA), or telephone (using the MoCA-BLIND version 7.1^30^). This affected 144 patient-visits in total (9.2 % of total patient-visits). The MoCA-Blind was scaled to the full MoCA score by dividing the MoCA-Blind score by the MoCA-Blind total score and multiplying by 30. Visuo-perceptual performance was assessed using a web-based platform that we described previously.^31^ This uses six tests of visuo-perception, of which four tests are most sensitive in PD^31^ and were used in the analysis presented here (see Supplement and^31^ for details of visuo-perceptual tests). We used performance on these visuo-perceptual tasks to divide people with PD into poor- and good-vision performers at baseline as follows: Patients scoring below the median score in two or more of the four visuo-perceptual tests were considered poor-vision PD; all other patients were considered good-vision PD^32^.

### Processing and analysis of genetic samples

DNA was extracted from whole blood. SNVs were genotyped using the Illumina NeuroChip Array (Infinium Core-24 Kit, n.d.) utilising Illumina’s standard protocol. NeuroChip incorporates a 306,670 variant tagging backbone (Infinium HumanCore-24 Exome) in addition to 179,467 custom variants linked to neurological disease.

*APOE* genotypes were inferred from the imputed genotypes of rs7412 and rs429358 variants (<u>10.1038/s41582-024-00988-2</u>). Patients with at least one *APOE4* allele were classified as *APOE4* positive. 18 patients had two *APOE4* alleles, which was too small a group to be considered separately. Participants with PD were divided into four groups based on their vision scores at baseline (poor-vision vs good-vision) and *APOE4* status (positive or negative). *GBA1* variants were identified from the imputed Neurochip data. E365K and T408M were specifically examined due to their association with cognitive decline^24,33,34^. Patients were marked *GBA1* positive if they had either of these variants. There were no patients with both variants. PD participants were similarly divided into four groups based on vision scores and *GBA1* variant positivity or negativity. Patients were classified as positive for *RIMS1* with at least one allele positive for the rs12528068 variant, and as *LRP1B* positive with at least one allele with the rs80306347 variant.

Polygenic risk scores (PRSs) were generated with PLINK v2 and v1.9,^35^ using GWAS studies for PD^36^, and AD.^37^ Prior to imputation, SNPs with an allele frequency of <0.01 were removed to avoid false positives. Exclusions for pre-imputation quality control included: sex mismatch, heterozygosity ±3SD from the mean, more than 5% missingness rate, minor allele frequency less than 0.05, or Hardy-Weinberg equilibrium exact test *p-*value greater than 0.0001. Imputation was performed using the Michigan Imputation server, using the Haplotype Reference Consortium version 1.1. Post imputation, quality control was performed prior to calculating PRS. PRS for PD and AD were divided into high vs low, using a median split, and for each of these, patients were divided into four groups, based on baseline visual performance (good- vs poor-vision PD) and PRS (low vs high, for both PD and AD). To examine combined effects of PD and AD-PRS, we also analysed a combined PD and AD-PRS group. This was generated by designating those below median in both PD and AD-PRS as negative (low score on both), and all others as positive (high PRS for PD or AD, or on both).

### Statistical Analyses

For demographic information, measures were compared between groups using two-tailed Welch’s t-tests or Mann-Whitney tests for non-normally distributed data, and Chi-squared test for categorical data. Survival curves were generated using the Kaplan-Meier method to calculate survival probabilities for data visualisation. Cox proportional hazard models were used to calculate a non-parametric estimate of survival times and probabilities adjusted for covariates including age at baseline and sex. These were calculated separately for visual performance, and for genetic groups. Progression to PD-MCI was defined when MoCA dropped below 25^29,38,39^ and subsequently remained ≤25 (as in previous work^16^). Progression to dementia was considered separately, using sustained MoCA≤21^39^. Time-to-event was defined as the date from baseline that MoCA became equal to or below the PD-MCI or PD dementia threshold, provided it stayed below the threshold at the following testing time. Data were right censored for participants who withdrew or were lost to follow-up. If there were no events in a group for the Cox proportional hazards model, the lower bound of the 95% confidence interval was used as the hazard ratio. Median follow-up was computed with the reverse Kaplan-Meier method. The survfit and survminer packages for R were used for all survival analysis. P<0.05 was accepted as threshold for statistical significance for demographic data, with false-discovery rate (FDR) correction for study comparisons. All data are presented as mean±standard deviation (SD) unless stated otherwise. Statistical analysis was carried out in R v4.3.2 (https://www.r-project.org/). Graphs were generated with ggplot2 R package.

Sample size requirements for a hypothetical disease modifying trial were calculated using simulation-based power calculations of subject-level longitudinal slopes equivalent to testing the treatment*time interaction under a linear mixed-effects model. Cognitive outcomes were modelled as linear change in MOCA scores over time, as measured at baseline, 18-months and 36-months. Between subject variability and within-subject residual variance components were derived empirically from our cohort. Simulated datasets reflected a planned trial design with assessments at baseline, 18- and 36-months. Treatment effects were specified as proportional reductions in the rates of cognitive decline. Subject-specific linear slopes were estimated from simulated longitudinal data and compared between treatment and control groups using two-sample tests. For each candidate sample size, 500 simulated trials were generated, with refitting of the mixed effects model. The minimum sample size per arm achieving 80% power with statistical significance α=0.05 is reported.

## Results

### Participant characteristics

587 patients with PD met selection criteria for the study. 137 did not complete follow-up visits after baseline tests were performed, including 28 patients who were excluded from analysis due to missing data from online vision tests at baseline (Figure 1). This left 450 people with PD included in the main analyses for this study (see Table 1 for demographic information). Mean age was 71.8±7.6 years. Mean disease duration at time of recruitment was 3.7±2.2 years. Mean MoCA at baseline was 27.7±1.6. 218 participants were classed as poor-vision PD at baseline, based on performance on visual tasks. Poor-vision PD participants were older compared with good-vision PD (74.2±7.0 vs 69.5±7.4 years, t=6.92, *p*<.0001), with a similar sex distribution (68% vs 66% males, χ²=0.2, *p*=.89), and slightly lower MoCA at baseline (27.3±1.6 vs 28.0±1.6, t=5.1 *p=*.01) (Table 1). They had slightly shorter disease duration of 3.3±2.1 years at baseline, compared to 4.0±2.3 years in good-vision PD participants (t=2.59, *p*=0.01). Mean follow-up duration was 32.7±12.3 months.

### PD patients with poor-vision have higher rates of progression to PD-MCI

PD patients with poor-vision had a higher risk of progression to PD-MCI compared to PD patients with good-vision, even after adjusting for age and sex. (HR=2.34, 95% CI=1.58-3.48, *p*FDR=0.00062, Fig 2A, Table 2). We found a similar effect for dementia, although due to lower numbers converting to dementia, this did not reach significance after FDR correction. (HR=4.25, 95% CI 1.40-12.9, *p*FDR*=*0.088), adjusted for age and sex (Fig 2B).

**Figure 2.**
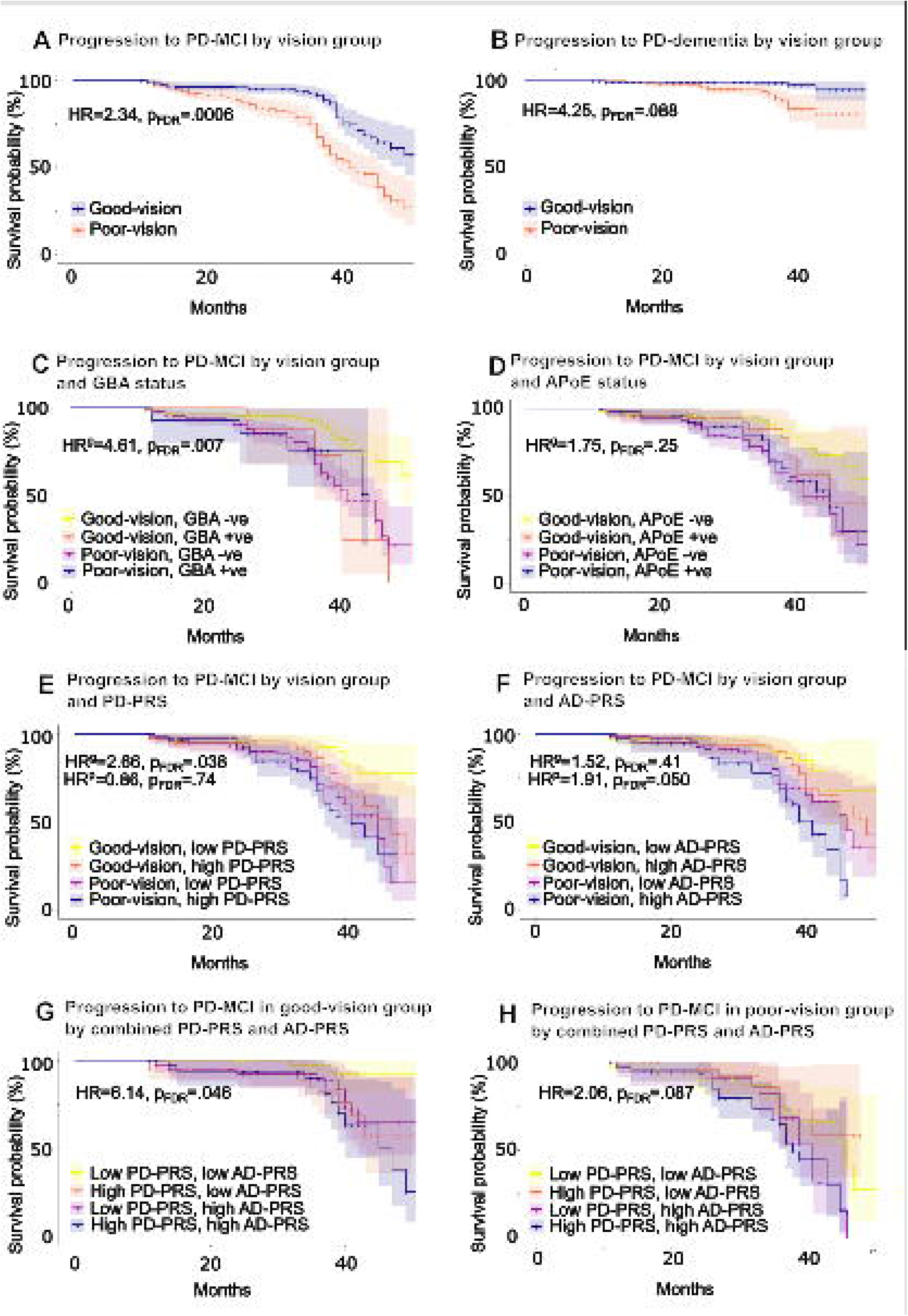
Survival curves for progression to mild cognitive impairment in PD (PD-MCI) stratifying by visual performance and *GBA* and *APOE*. Kaplan-Meier plots illustrating: A) Progression to PD-MCI for good and poor-vision PD B) Progression to PD-dementia for good and poor-vision PD C) Progression to PD-MCI for PD patients, divided by vision (good- versus poor-); and by *GBA1* risk variant status (positive vs negative for E365K or T408M). D) Progression to PD-MCI for PD patients, divided by vision (good- versus poor-); and by *APOE4* status (positive vs negative). E) Progression to PD-MCI for PD patients, divided by vision (good- versus poor-); and by PD-PRS status (low vs high). F) Progression to PD-MCI for PD patients, divided by vision (good- versus poor); and by AD-PRS status (low vs high). G) Progression to PD-MCI for PD patients with good vision, divided by combined PD-PRS and AD-PRS status (both low; high PD-PRS, high AD-PRS; both high). H) Progression to PD-MCI for PD patients with poor vision, divided by combined PD-PRS and AD-PRS status (both low; high PD-PRS, high AD-PRS; both high). PD-MCI was defined using MoCA score <=25. Timeline is in months. Shaded area represents 95% CIs. Log rank score was calculated using Cox’s proportional hazards, with age- and sex-correction. AD, Alzheimer’s disease; HR, Hazard Ratio; HR^g^, hazard ratio within good-vision groups; HR^p^, hazard ratio within poor vision groups; PD, Parkinson’s disease; PRS, polygenic risk score.

### Effect of *GBA1* risk variants and *APOE4* polymorphism on progression to PD-MCI, and interaction with vision status

10 patients carried the *GBA1* E365K risk variant and 14 carried a *GBA1* T408M variant. The total number of cases with either *GBA1* variant (E365K or T408M) was small (n=24), and these were not associated with a significantly higher overall rate of progression to PD-MCI (HR=1.68, CI=0.84-3.36, *p*FDR=0.24774). There was only 1 conversion to dementia event in this group which was too small to examine formally.

We found a differential effect of *GBA1* according to vision-status. For PD patients with good-vision, carrying either *GBA1* variant was associated with increased risk of progression to PD-MCI over the time period (HR=4.61, CI=1.73-12.28, *p*FDR=0.0068, age and sex-corrected. Fig. 2C, Table 2). However, in patients with poor vision, *GBA1* variant status did not affect progression to PD-MCI (HR=1.27, CI=0.46-3.52, *p*FDR=0.77, Supplementary Table 2) or PD dementia (Supplementary Fig. 1A and Supplementary Table 1).

110 patients carried the *APOE4* polymorphism. This was not associated with an increased risk of progression to PD-MCI (HR=1.178, CI=0.75-1.85, *p*FDR=0.72), or dementia (HR=1.17, CI=0.39-3.49, *p*FDR=0.97, Supplementary Fig. 1B). We also found no differential effect of *APOE4* polymorphism according to vision status. Carrying the *APOE4* polymorphism did not affect progression to PD-MCI for patients with poor-vision (HR=1.25, CI=0.71-2.20, *p*FDR=0.60, Supplementary Table 2) or good-vision (HR=1.75, CI=0.81-3.76, *p*FDR=0.25, Fig. 1D, Table 2); and no differences were seen for progression to dementia (Supplementary Fig 1B, Supplementary Table 1). We also examined effects of *LRP1B* and *RIMS1* but did not find significant effects on progression to PD-MCI or dementia (Supplementary Fig.3 and 4).

### Effect of Polygenic Risk Score on progression to PD-MCI and interaction with vision

325 patients were included in the PRS analysis. High PD-PRS was not associated with increased progression to PD-MCI (HR=1.64, CI=1.06-2.54, *p*FDR=0.058), nor high AD-PRS (HR=1.65, CI=1.060-2.56, *p*FDR=0.057). Neither were associated with progression to dementia (PD-PRS HR=5.10, CI=1.40-18.65, *p*FDR=0.11, AD-PRS HR=3.73, CI=1.038-13.42, *p*FDR=0.15).

For the differential effect of PD-PRS stratified by vision status, PD-PRS status did not affect progression to PD-MCI in poor-vision PD patients (HR=0.86, CI=0.50-1.47, *p*FDR=0.74, Supplementary Table 2). However, having a high PD-PRS was associated with increased progression to PD-MCI in good-vision PD patients (HR=2.66, CI=1.21-5.81, *p*FDR=0.038, Table 2, Fig.2E). PD-PRS had no effect on progression to dementia in good or poor vision patients, likely due to low numbers of conversions to dementia in the cohort.

On examining the differential effect of AD-PRS stratified by vision status, we found a similarly negative effect on progression in patients with high AD-PRS, which was seen exclusively in the poor-vision PD patients. We found no significant difference in progression to PD-MCI between high and low AD-PRS in good-vision PD (HR=1.52, CI=0.71-3.25, *p*FDR=0.41; Table 2); but in poor-vision PD patients, having a high AD-PRS was linked with higher progression to PD-MCI (HR=1.91, CI=1.10-3.32, *p*FDR=0.04999; Fig. 1F, Supplementary Table 2). This effect was not observed with dementia-free survival, likely due to a smaller number of conversion events (Supplementary Table 1, Supplementary Fig. 2).

We also examined the combined effects of PD- and AD-PRS. Having both high PD- and AD-PRS was associated with overall greater progression to PD-MCI (HR=2.02, CI=1.16-3.51, *p*FDR=0.032). On examining the differential effect of both high PD- and AD-PRS, stratified based on vision status, we found that patients with good-vision and both high PD- and AD-PRS demonstrated markedly higher progression to PD-MCI compared to patients with low PD- and AD-PRS scores (HR=6.14, CI=1.36-27.83, *p*FDR=0.046, Fig.2G). In patients with poor-vision, the combination of high PD- and AD-PRS scores was also associated with increased progression to PD-MCI than in those with low PD- and AD-PRS (HR=2.06, CI=1.013-4.20, *p*FDR=0.087, Fig. 2H), although this did not survive correction (unadjusted *p*= 0.046).

### Effect of including visual and genetic information on sample sizes for clinical trials

Our simulations revealed that for an agent slowing cognitive decline by 30%, without stratification, a sample size of n=705 would be required. Genetic stratification selecting patients with PD with both high PD- and AD-PRS, could reduce the sample size to n=285. Stratification using our visual test reduced the sample size to n=285; and stratifying by combining both poor vision and high-PD- and AD-PRS reduced the required sample size to n=160 (Table 3).

## Discussion

In this large multi-centre longitudinal cohort study with mean follow-up 32.7 months, we investigated the predictive effects of visual function on cognitive outcomes in PD, and how these combine with genetic factors. We found that poor performance on visual tests in PD predicted higher progression to PD-MCI and dementia. We further demonstrated an additive effect of genetic factors on visual performance. Specifically, we found that in PD patients with good vision, carrying either E326K or T408M *GBA1* variants had even greater PD-MCI progression. We also found that having a high PD-PRS increased PD-MCI progression in PD patients in the protected good-vision group; and that high AD-PRS was associated with greater PD-MCI progression in poor-vision PD patients. These effects on progression to PD-MCI of PD- and AD-PRS were even stronger in combination, with a negative effect on progression for combined high PD- and AD-PRS; and a profoundly protective effect on survival for patients with both low PD- and AD-PRS. Our simulations revealed that using vision and genetics to enrich clinical trials by those patients most likely to develop dementia, can significantly reduce the required sample sizes.

Our findings of poor visual function predicting poor outcomes in PD support previous work in smaller cohorts. A prospective study of 142 newly-diagnosed PD patients showed that those making errors copying pentagons were at double the risk of developing dementia within 10 years^2^; and in a prospective study of 80 patients, impaired colour vision was linked with an odds ratio of 3.3 for dementia after follow-up^15^. More recently, we showed in 100 people with PD followed-up for 3 years, that poor baseline performance on visual tests was linked with higher risk of developing dementia and other poor outcomes, including death^16^. This association between poor vision and poor outcomes in PD has also been found in large population studies in the US and South Korea^17,40^, although, those studies were retrospectively analysed. We now show in a large cohort of prospectively-examined patients, that visual function predicts cognitive outcomes in PD, even when accounting for age and sex.

The link between *GBA1* risk variants and cognitive decline in PD is well-established^23^. Here, we now demonstrate that genetic factors have an additive effect to clinical predictors such as visual dysfunction on risk of cognitive decline in PD. An association between PD-PRS and cognitive decline was previously found in a smaller study of 285 patients, with mean follow-up 5.3 years ^27^, but how this interacts with AD-PRS in a prospective cohort had not yet been examined.

We found no significant effect for *APOE4* carriers and higher conversion to PD dementia or PD-MCI. Whilst several previous studies have shown a link between *APOE4* and increased risk of PD dementia.^19,23,25,41,42^, other studies have not found this association.^21,22,43^. It is likely that these differences arise due to sampling effects between populations, and that an association with *APOE4* does exist, but only in some populations.

### Strengths and limitations

We present a large, prospectively-collected study examining visual function and global cognition with genetics in PD. Patients were recruited from PD clinics with light-touch assessments, designed to widen participation and reflect a real-world PD dataset, rather than selection for fitter patients, required for attending extended research visits. However, we lacked detailed cognitive outcomes, motor MDS-UPDRS scores, and information on other factors contributing to increased risk of PD dementia, including anxiety and depression, REM sleep behaviour disorder, ethnicity and socio-economic status.

During the follow-up period, some patients were unable to attend in person for follow-up, due to the COVID-19 pandemic. We used video or telephone visits to perform MoCA assessments, and adjusted telephone MoCA scores to account for this. As cognitive change in PD disproportionately affects visual components of the MoCA^43^, this is likely to reduce the sensitivity of our results by artificially elevating some scores. Our finding of an association between poor visual function and cognitive change over time, despite reduced sensitivity of phone or video-share MoCA, suggests that these relationships are robust.

There was loss to follow-up, as expected in a real-world longitudinal study, with greater loss in patients with higher levels of frailty or cognitive involvement. Despite lower numbers at follow-up, with attrition in the most affected group, we were still able to detect associations between visual performance, genetic factors and cognitive outcomes in PD.

## Conclusions

We showed in a large prospective longitudinal study that patients performing poorly on visual function tests are at increased risk of developing cognitive decline, and that this acts in combination with genetic factors of *GBA1* risk variants and PD- and AD-PRS. The data reported in our study can improve the efficiency of clinic trials for PD dementia by using visual and genetic factors to identify those patients at highest risk for PD dementia, who are likely to progress to PD dementia most rapidly. It can also be combined with other clinical and biomarker information to improve prognostic information for patients with PD in the clinical setting.

## Data Availability

Anonymised data will be shared by request from a qualified investigator.

## Abbreviations

AD: Alzheimer’s disease
GWAS: genome wide association study
MCI: Mild cognitive impairment
MoCA: Montreal Cognitive Examination
PD: Parkinson’s disease
PDD: Parkinson’s disease dementia
PRS: polygenic risk score

## Funding

RSW was supported by a Wellcome Clinical Research Career Development Fellowship (205167/Z/16/Z) and by a Wellcome Career Development Award (225263/Z/22/Z). This work was also funded by grants from UCLH Biomedical Research Centre Grant (BRC302/NS/RW/101410).

## Competing interests

RSW has received honoraria from GE Healthcare, Bial, Omnix Pharma, the Shirley Ryan Ability Lab and Britannia, and consultancy from Therakind and Accenture.

## Data Sharing statement

Data available: yes

Data types: deidentified participant data, data dictionary.

How to access data: anonymised data will be shared by request from a qualified investigator.

When available: after publication

---

**Table 1**

Demographic information for patients with Parkinson’s disease (PD), and for PD patients with good-vision and poor-vision, defined using web-based test of visuo-perception. MoCA, Montreal Cognitive Assessment; PD, Parkinson’s disease

**Table 2**

Cox-proportional hazard model summary statistics for PD patients across all analysis groups, comparing good and poor vision adjusted for age, sex, and genetic factors, and using good-vision patients as the reference group. AD, Alzheimer’s disease; MCI, mild cognitive impairment; PD, Parkinson’s disease; PRS, polygenic risk score.

**Table 3**

Sample size estimates based on simulations for an intervention slowing cognitive decline. AD-PRS, Alzheimer’s Disease polygenic risk score; MoCA, Montreal Cognitive Assessment; PD, Parkinson’s disease; PD-PRS, Parkinson’s disease polygenic risk score.

## Notes

### Author Declarations

Queen Square Research Ethics Committee of University College London gave ethical approval for this work

## References

1. Ray Dorsey E, Elbaz A, Nichols E, et al. Global, regional, and national burden of Parkinson’s disease, 1990–2016: a systematic analysis for the Global Burden of Disease Study 2016. Lancet Neurol. 2018;17(11):939. doi:10.1016/S1474-4422(18)30295-3

2. Williams-Gray CH, Mason SL, Evans JR, et al. The CamPaIGN study of Parkinson’s disease: 10-year outlook in an incident population-based cohort. J Neurol Neurosurg Psychiatry. 2013;84(11):1258–1264. doi:10.1136/JNNP-2013-305277

3. Poewe W. Non-motor symptoms in Parkinson’s disease. Eur J Neurol. Published online 2008. doi:10.1111/j.1468-1331.2008.02056.x

4. Levy G, Tang MX, Louis ED, et al. The association of incident dementia with mortality in PD. Neurology. Published online 2002. doi:10.1212/01.WNL.0000036610.36834.E0

5. Xu Y, Yang J, Shang H. Meta-analysis of risk factors for Parkinson’s disease dementia. Transl Neurodegener. Published online 2016. doi:10.1186/s40035-016-0058-0

6. Simuni T, Chahine LM, Poston K, et al. A biological definition of neuronal α-synuclein disease: towards an integrated staging system for research. Lancet Neurol. Published online 2024. doi:10.1016/S1474-4422(23)00405-2

7. Höglinger GU, Adler CH, Berg D, et al. A biological classification of Parkinson’s disease: the SynNeurGe research diagnostic criteria. Lancet Neurol. Published online 2024. doi:10.1016/S1474-4422(23)00404-0

8. Irwin DJ, Grossman M, Weintraub D, et al. Neuropathological and genetic correlates of survival and dementia onset in synucleinopathies: a retrospective analysis. Lancet Neurol. Published online 2017. doi:10.1016/S1474-4422(16)30291-5

9. Toledo JB, Abdelnour C, Weil RS, et al. Dementia with Lewy bodies: Impact of co-pathologies and implications for clinical trial design. Alzheimer’s Dement. Published online 2023. doi:10.1002/alz.12814

10. Coughlin DG, Hurtig HI, Irwin DJ. Pathological Influences on Clinical Heterogeneity in Lewy Body Diseases. Mov Disord. Published online 2020. doi:10.1002/mds.27867

11. Vijiaratnam N, Girges C, Auld G, et al. Exenatide once weekly over 2 years as a potential disease-modifying treatment for Parkinson’s disease: Protocol for a multicentre, randomised, double blind, parallel group, placebo controlled, phase 3 trial: The Exenatide-PD3’ study. BMJ Open. Published online 2021. doi:10.1136/bmjopen-2020-047993

12. Evans JR, Mason SL, Williams-Gray CH, et al. The natural history of treated Parkinson’s disease in an incident, community based cohort. J Neurol Neurosurg Psychiatry. Published online 2011. doi:10.1136/jnnp.2011.240366

13. Schrag A, Siddiqui UF, Anastasiou Z, Weintraub D, Schott JM. Clinical variables and biomarkers in prediction of cognitive impairment in patients with newly diagnosed Parkinson’s disease: a cohort study. Lancet Neurol. Published online 2017. doi:10.1016/S1474-4422(16)30328-3

14. Liu G, Locascio JJ, Corvol JC, et al. Prediction of cognition in Parkinson’s disease with a clinical–genetic score: a longitudinal analysis of nine cohorts. Lancet Neurol. Published online 2017. doi:10.1016/S1474-4422(17)30122-9

15. Anang JBM, Gagnon JF, Bertrand JA, et al. Predictors of dementia in Parkinson disease: A prospective cohort study. Neurology. Published online 2014. doi:10.1212/WNL.0000000000000842

16. Hannaway N, Zarkali A, Leyland LA, et al. Visual dysfunction is a better predictor than retinal thickness for dementia in Parkinson’s disease. J Neurol Neurosurg Psychiatry. Published online 2023. doi:10.1136/jnnp-2023-331083

17. Hamedani AG, Abraham DS, Maguire MG, Willis AW. Visual Impairment Is More Common in Parkinson’s Disease and Is a Risk Factor for Poor Health Outcomes. Mov Disord. Published online 2020. doi:10.1002/mds.28182

18. Mata IF, Leverenz JB, Weintraub D, et al. GBA Variants are associated with a distinct pattern of cognitive deficits in Parkinson’s disease. Mov Disord. Published online 2016. doi:10.1002/mds.26359

19. Mata IF, Leverenz JB, Weintraub D, et al. APOE, MAPT, and SNCA genes and cognitive performance in Parkinson disease. JAMA Neurol. Published online 2014. doi:10.1001/jamaneurol.2014.1455

20. Tropea TF, Xie SX, Rick J, et al. APOE, thought disorder, and SPARE-AD predict cognitive decline in established Parkinson’s disease. Mov Disord. Published online 2018. doi:10.1002/mds.27204

21. Federoff M, Jimenez-Rolando B, Nalls MA, Singleton AB. A large study reveals no association between APOE and Parkinson’s disease. Neurobiol Dis. Published online 2012. doi:10.1016/j.nbd.2012.02.002

22. Mengel D, Dams J, Ziemek J, et al. Apolipoprotein E ε4 does not affect cognitive performance in patients with Parkinson’s disease. Park Relat Disord. Published online 2016. doi:10.1016/j.parkreldis.2016.04.013

23. Szwedo AA, Dalen I, Pedersen KF, et al. GBA and APOE Impact Cognitive Decline in Parkinson’s Disease: A 10-Year Population-Based Study. Mov Disord. Published online 2022. doi:10.1002/mds.28932

24. Liu G, Peng J, Liao Z, et al. Genome-wide survival study identifies a novel synaptic locus and polygenic score for cognitive progression in Parkinson’s disease. Nat Genet. Published online 2021. doi:10.1038/s41588-021-00847-6

25. Real R, Martinez-Carrasco A, Reynolds RH, et al. Association between the LRP1B and APOE loci and the development of Parkinson’s disease dementia. Brain. Published online 2023. doi:10.1093/brain/awac414

26. Lewis CM, Vassos E. Polygenic risk scores: From research tools to clinical instruments. Genome Med. Published online 2020. doi:10.1186/s13073-020-00742-5

27. Paul KC, Schulz J, Bronstein JM, Lill CM, Ritz BR. Association of polygenic risk score with cognitive decline and motor progression in Parkinson disease. JAMA Neurol. Published online 2018. doi:10.1001/jamaneurol.2017.4206

28. Postuma RB, Berg D, Stern M, et al. MDS clinical diagnostic criteria for Parkinson’s disease. Mov Disord. Published online 2015. doi:10.1002/mds.26424

29. Nasreddine ZS, Phillips NA, Bédirian V, et al. The Montreal Cognitive Assessment, MoCA: A brief screening tool for mild cognitive impairment. J Am Geriatr Soc. Published online 2005. doi:10.1111/j.1532-5415.2005.53221.x

30. Pendlebury ST, Welch SJV, Cuthbertson FC, Mariz J, Mehta Z, Rothwell PM. Telephone assessment of cognition after transient ischemic attack and stroke: Modified telephone interview of cognitive status and telephone montreal cognitive assessment versus face-to-face montreal cognitive assessment and neuropsychological battery. Stroke. Published online 2013. doi:10.1161/STROKEAHA.112.673384

31. Weil RS, Schwarzkopf DS, Bahrami B, et al. Assessing cognitive dysfunction in Parkinson’s disease: An online tool to detect visuo-perceptual deficits. Mov Disord. Published online 2018. doi:10.1002/mds.27311

32. Zarkali A, McColgan P, Leyland LA, Lees AJ, Rees G, Weil RS. Fiber-specific white matter reductions in Parkinson hallucinations and visual dysfunction. Neurology. Published online 2020. doi:10.1212/WNL.0000000000009014

33. Iwaki H, Blauwendraat C, Leonard HL, et al. Genetic risk of Parkinson disease and progression: An analysis of 13 longitudinal cohorts. Neurol Genet. Published online 2019. doi:10.1212/NXG.0000000000000348

34. Davis MY, Johnson CO, Leverenz JB, et al. Association of GBA mutations and the E326K polymorphism with motor and cognitive progression in parkinson disease. JAMA Neurol. Published online 2016. doi:10.1001/jamaneurol.2016.2245

35. Purcell S, Neale B, Todd-Brown K, et al. PLINK: A tool set for whole-genome association and population-based linkage analyses. Am J Hum Genet. Published online 2007. doi:10.1086/519795

36. Nalls MA, Blauwendraat C, Vallerga CL, et al. Identification of novel risk loci, causal insights, and heritable risk for Parkinson’s disease: a meta-analysis of genome-wide association studies. Lancet Neurol. Published online 2019. doi:10.1016/S1474-4422(19)30320-5

37. Kunkle BW, Grenier-Boley B, Sims R, et al. Genetic meta-analysis of diagnosed Alzheimer’s disease identifies new risk loci and implicates Aβ, tau, immunity and lipid processing. Nat Genet. Published online 2019. doi:10.1038/s41588-019-0358-2

38. Thomann AE, Berres M, Goettel N, Steiner LA, Monsch AU. Enhanced diagnostic accuracy for neurocognitive disorders: A revised cut-off approach for the Montreal Cognitive Assessment. Alzheimer’s Res Ther. Published online 2020. doi:10.1186/s13195-020-00603-8

39. Dalrymple-Alford JC, MacAskill MR, Nakas CT, et al. The MoCA: Well-suited screen for cognitive impairment in Parkinson disease. Neurology. Published online 2010. doi:10.1212/WNL.0b013e3181fc29c9

40. Han G, Han J, Han K, Youn J, Chung TY, Lim DH. Visual Acuity and Development of Parkinson’s Disease: A Nationwide Cohort Study. Mov Disord. Published online 2020. doi:10.1002/mds.28184

41. Pankratz N, Byder L, Halter C, et al. Presence of an APOE4 allele results in significantly earlier onset of Parkinson’s disease and a higher risk with dementia. Mov Disord. Published online 2006. doi:10.1002/mds.20663

42. Tunold JA, Geut H, Rozemuller JMA, et al. APOE and MAPT Are Associated With Dementia in Neuropathologically Confirmed Parkinson’s Disease. Front Neurol. Published online 2021. doi:10.3389/fneur.2021.631145

43. Williams-Gray CH, Mason SL, Evans JR, et al. The CamPaIGN study of Parkinson’s disease: 10-year outlook in an incident population-based cohort. J Neurol Neurosurg Psychiatry. Published online 2013. doi:10.1136/jnnp-2013-305277

