## Supplementary material for "Clinical and genetic predictors of dementia in Parkinson’s disease": Table 1

**Table 1. Demographic and clinical information of all patients with Parkinson’s disease (PD) and patients separated into good- and poor- vision**

|  | **Parkinson’s Disease (PD)** | **PD Good- vision** | **PD Poor-vision** | **Test Statistic (good vs poor vision)** | **p-value** |
| --- | --- | --- | --- | --- | --- |
| *n* | 450 | 232 | 218 |  |  |
| Baseline Age (years) | 71.78 (7.59) | 69.50 (7.44) | 74.21 (6.98) | t=6.92 | **<.0001** |
| Sex  (% Male / Female) | 67/33 | 66/34 | 68/32 | χ²=0.2 | .89 |
| Baseline disease duration (years) | 3.65 (2.21) | 3.95 (2.30) | 3.34 (2.07) | t=2.59 | **.010** |
| Baseline MoCA | 27.67 (1.63) | 28.04 (1.60) | 27.28 (1.57) | t=5.05 | **<.0001** |
| Hallucinations  (% with) * | 7.2% | 4.0% | 10.7% | χ²=6.94 | **.031** |
| ApoE-E4  (% positive / negative) | 27/73 | 23/67 | 32/68 | χ²=2.56 | .11 |
| GBA  (% positive / negative) | 7/93 | 6/94 | 8/92 | χ²=0.57 | .45 |
| PD-PRS | -3.53 (0.98) | -3.57 (0.95) | -3.50 (1.02) | t=0.64 | .52 |
| AD-PRS | -0.04 (1.12) | -0.04 (1.25) | -0.04 (0.97) | t=0.05 | .96 |
| Duration of follow-up (months) | 32.50 (11.89) | 33.60 (12.02) | 31.56 (11.85) | t=1.91 | .056 |
| PD-MCI during follow up (% with) | 27.8% | 18.1% | 38.1% | χ²=21.36 | **<.0001** |
| PDD during follow up (% with) | 6.7% | 2.2% | 7.3% | χ²=5.67 | **.017** |

All values are reported mean(standard deviation), apart from sex and hallucinations which are reported as percentages. MoCA = Montreal Cognitive Assessment. * Presence/absence of hallucinations reported for 367 participants

**Table 2. Models of Progression to Mild Cognitive Impairment**

| Model (comparator group) | Variable | HR (CI) | FDR corrected p-value |
| --- | --- | --- | --- |
| Vision Group + Age + Sex (Good vision) | | | |
|  | Poor vision | 2.34 (1.58 - 3.48) | **.0006** |
|  | Age | 1.05 (1.02 - 1.08) | **.002** |
|  | Sex | 0.99 (0.67 - 1.47) | .99 |
| GBA mutations/Vision Group + Age + Sex (Good vision / GBA negative) | | | |
|  | Good vision / GBA positive | 4.61 (1.73 – 12.28) | **.007** |
|  | Poor vision / GBA negative | 2.75 (1.68 – 4.51) | **.001** |
|  | Poor vision / GBA positive | 2.16 (0.74 – 6.32) | .25 |
|  | Age | 1.06 (1.03 – 1.10) | **.002** |
|  | Sex | 0.88 (0.55 – 1.41) | .76 |
| ApoE-E4 polymorphism/Vision Group + Age + Sex (Good vision / ApoE-E4 negative) | | | |
|  | Good vision / ApoE-E4 positive | 1.75 (0.81 – 3.76) | .25 |
|  | Poor vision / ApoE-E4 negative | 2.65 (1.50 – 4.67) | **.003** |
|  | Poor vision / ApoE-E4 positive | 2.12 (1.11 – 4.02) | **.050** |
|  | Age | 1.06 (1.02 – 1.10) | **.003** |
|  | Sex | 0.98 (0.60 – 1.58) | .98 |
| PD-PRS + Age + Sex (low PD-PRS) | | | |
|  | High PD-PRS | 1.64 (1.06 – 2.54) | .058 |
|  | Age | 1.08 (1.04 – 1.11) | **.0003** |
|  | Sex | 1.07 (0.66 – 1.74) | .90 |
| AD-PRS + Age + Sex (low AD-PRS) | | | |
|  | High AD-PRS | 1.65 (1.06 – 2.56) | .057 |
|  | Age | 1.08 (1.04 – 1.11) | **.0003** |
|  | Sex | 1.00 (0.62 – 1.61) | .99 |
| PD-PRS/Vision Group + Age + Sex (Good vision / low PD-PRS) | | | |
|  | Good vision / high PD-PRS | 2.66 (1.21 – 8.81) | **.038** |
|  | Poor vision / low PD-PRS | 3.18 (1.49 – 6.83) | **.009** |
|  | Poor vision / high PD-PRS | 3.72 (1.72 – 8.05) | **.003** |
|  | Age | 1.06 (1.03 – 1.10) | **.002** |
|  | Sex | 1.05 (0.64 = 1.70) | .94 |
| AD-PRS /Vision Group + Age + Sex (High vision /low AD-PRS) | | | |
|  | Good vision / high AD-PRS | 1.51 (0.71 – 3.25) | .41 |
|  | Poor vision / low AD-PRS | 1.78 (0.83 – 3.80) | .24 |
|  | Poor vision / high AD-PRS | 3.41 (1.65 – 7.01) | **.003** |
|  | Age | 1.07 (1.03 – 1.10) | .002 |
|  | Sex | 1.01 (-.62 – 1.63) | .99 |
| PD-PRS / AD-PRS + Age + Sex, in good vision group only (Low AD-PRS / Low PD-PRS) | | | |
|  | High AD-PRS / Low PD-PRS | 4.16 (0.87 – 19.84) | .14 |
|  | Low AD-PRS / High PD-PRS | 6.72 (1.40 – 32.18) | **.044** |
|  | High AD-PRS / High PD-PRS | 6.14 (1.36 – 27.83) | **.046** |
|  | Age | 1.05 (1.00 – 1.11) | .076 |
|  | Sex | 1.94 (0.75 – 4.99) | .26 |
| PD-PRS / AD-PRS + Age + Sex, in poor vision group only (Low AD-PRS / Low PD-PRS) | | | |
|  | High AD-PRS / Low PD-PRS | 1.70 (0.80 – 3.65) | .26 |
|  | Low AD-PRS / High PD-PRS | 0.69 (0.26 – 1.81) | .61 |
|  | High AD-PRS / High PD-PRS | 2.06 (1.01 – 4.20) | .087 |
|  | Age | 1.08 (1.02 - 1.13) | **.005** |
|  | Sex | 0.84 (0.47 – 1.52) | .74 |

AD-PRS = Alzheimer’s Disease polygenic risk score, ApoE-E4 = Apolipoprotein E-E4 , GBA = glucosylceramidase beta, HR = hazard ratio, PD-PRS = Parkinson’s disease polygenic risk score, SE = standard error

**Table 3. Sample size estimates based on simulations for an intervention slowing cognitive decline**

| **Patient stratification method** | **Annual Change in MoCA**  **mean (SD)** | **Agent showing ~10% slowing in cognitive decline** | **Agent showing ~20% slowing in cognitive decline** | **Agent showing ~30% slowing in cognitive decline** |
| --- | --- | --- | --- | --- |
| Unselected PD group | -0.240 (1.263) | n= 6425 | n= 1615 | n= 705 |
| Unselected PD group + PD PRS | -0.257 (1.324) | n= 5475 | n= 1295 | n= 630 |
| Unselected PD group + PD PRS + AD PRS | -0.381 (1.269) | n= 2575 | n= 650 | n= 285 |
| PD poor vision | -0.425 (1.468) | n= 2365 | n= 650 | n= 285 |
| PD poor vision + AD PRS | -0.499 (1.534) | n= 1675 | n= 470 | n= 205 |
| PD poor vision + AD PRS + PD PRS | -0.538 (1.531) | n= 1585 | n= 345 | n= 160 |

AD-PRS = Alzheimer’s Disease polygenic risk score, MoCA = Montreal Cognitive Assessment, PD = Parkinson’s disease, PD-PRS = Parkinson’s disease polygenic risk score
