## Supplementary Materials for "Clinical and genetic predictors of dementia in Parkinson’s disease"

**Supplementary Methods**

**Details on visual tests**

Participants accessed the visual tests via a web-based platform, as described previously^1^. In brief, four tests were included in the study described here: object invariance, peripheral object recognition, hidden figures, and mental rotation. Object invariance is a test using skewed images of cats and dogs. Participants were shown skewed images for 450 milliseconds, followed by a choice screen (3000 milliseconds, 24 trials). Peripheral object recognition involved participants viewing images of animals in pairs, one centrally, and one in the periphery, with side of presentation pseudorandomised. The images were either matched or different. In order to ensure patients fixated the central images and viewed the peripheral image in the periphery, the images were unequal in size, with the central image much smaller. Patients underwent 24 trials. Hidden figures involved viewing an image with 22 horses, of which 7 were unhidden, and the remaining 15 were hidden in the surrounding scenery. Patients were instructed to click on all the horses they could see. Number of horses detected, incorrect selections, repeat selections and total time were recorded. Mental rotation involved viewing a reference 5 x 5 grid, below which were presented three test grids. Within each grid were 4 blue circles in pseudorandom positions. The test grids were rotated 180˚ or ±90˚. In 2 of the test grids, one of the blue circles was moved to another position, adjacent to its equivalent position in the reference gird. The task was to select the rotated grid that matched the reference grid. Patients performed 24 trials. Missed trials were removed.

**Sample size calculations for milestones of conversion to PD-MCI or PD dementia**

We estimated required sample sizes for the time-to-endpoint, with cognitive endpoints being either PD-MCI or PD dementia using a Monte Carlo simulation framework based on empirical time-to-event data from our cohort. For each enrichment strategy (overall unselected PD population, PD with poor vision, and combinations with polygenic risk scores), we first parametrised the baseline survival distribution by fitting a Weibull model to observed survival times and event indicators.

The fitted Weibull model’s shape and scale parameters were then used to generate synthetic control survival times^2^, while treatment arm survival times were generated under a range of assumed hazard ratios (HRs) reflecting putative therapeutic effects (10%, 20% and 30% delay in the onset of MCI/dementia). Administrative censoring at a fixed follow-up time of 36 months, similar to our cohort, was applied in each simulation, and right-censored survival times were analysed using the log-rank test to compare the two arms. For each candidate sample size per arm, 500 simulations were run, and the proportion of log-rank tests achieving statistical significance at α=0.05 was recorded as estimated power. The smallest sample size per arm achieving ≥80 % power was selected as the required sample size for that strategy. This simulation-based approach accommodates realistic censoring patterns, non-exponential hazards, and complex subgroup structures that are not captured by simple closed-form event proportion calculations.

**Supplementary Table 1. Models of Survival Time for progression to Parkinson’s dementia**

| Model (comparator group) | Variable | HR (CI) | FDR corrected p-value |
| --- | --- | --- | --- |
| Vision Group + Age + Sex (Good vision) | | | |
|  | Poor vision | 4.25 (1.40 – 12.89) | .11 |
|  | Age | 1.12 (1.03 – 1.20) | .11 |
|  | Sex | 0.99 (0.40 – 2.43) | .99 |
| *GBA* mutations/Vision Group + Age + Sex (Good vision / *GBA* negative) | | | |
|  | Good vision / *GBA* positive | 0.00 (0 – infinite) | .99 |
|  | Poor vision / *GBA* negative | 4.00 (1.11 – 14.43) | .13 |
|  | Poor vision / *GBA* positive | 3.57 (0.37 – 34.63) | .63 |
|  | Age | 1.13 (1.03 – 1.24) | .11 |
|  | Sex | 0.85 (0.29 – 2.48) | .99 |
| *APOE*-E4 polymorphism/Vision Group + Age + Sex (Good vision / ApoE-E4 negative) | | | |
|  | Good vision / APOE-E4 positive | 0.00 (0 – infinite) | .99 |
|  | Poor vision / APOE-E4 negative | 2.81 (0.71 – 11.18) | .38 |
|  | Poor vision / APOE-E4 positive | 3.26 (0.76 – 13.95) | .31 |
|  | Age | 1.10 (1.01 – 1.21) | .13 |
|  | Sex | 0.99 (0.31 – 3.19) | .99 |
| PD-PRS + Age + Sex (low PD-PRS) | | | |
|  | High PD-PRS | 5.10 (1.40 – 18.65) | .11 |
|  | Age | 1.13 (1.04 – 1.23) | .11 |
|  | Sex | 1.12 (0.;34 – 3.65) | .99 |
| AD-PRS + Age + Sex (low AD-PRS) | | | |
|  | High AD-PRS | 3.73 (1.04 – 13.42) | .15 |
|  | Age | 1.12 (1.03 – 1.22) | .11 |
|  | Sex | 0.87 (0.27 – 2.81) | .99 |
| PD-PRS/Vision Group + Age + Sex (Good vision / low PD-PRS) | | | |
|  | Good vision / high PD-PRS | 1.27 x10^8^ (0 – infinite) | .99 |
|  | Poor vision / low PD-PRS | 9.17 x10^7^ (0 – infinite) | .99 |
|  | Poor vision / high PD-PRS | 3.42 x10^8^ (0 – infinite) | .99 |
|  | Age | 1.11 (1.01 – 1.22) | .13 |
|  | Sex | 1.15 (0.36 – 3.74) | .99 |
| AD-PRS /Vision Group + Age + Sex (Good vision /low AD-PRS) | | | |
|  | Good vision / high AD-PRS | 1.42 (0.13 – 15.80) | .99 |
|  | Poor vision / low AD-PRS | 1.28 (0.11 – 14.67) | .99 |
|  | Poor vision / high AD-PRS | 8.13 (1.01 – 65.78) | .16 |
|  | Age | 1.11 (1.01 – 1.22) | .13 |
|  | Sex | 1.15 (0.36 – 3.74) | .99 |
| PD-PRS / AD-PRS + Age + Sex, in good vision group only (Low AD-PRS / Low PD-PRS) | | | |
|  | High AD-PRS / Low PD-PRS | 1.06 x10^10^ (0 – infinite) | .99 |
|  | Low AD-PRS / High PD-PRS | 1.68 (0 – infinite) | .99 |
|  | High AD-PRS / High PD-PRS | 5.13 x10^9^ (0 – infinite) | .99 |
|  | Age | 1.26 (1.03 – 1.54) | .13 |
|  | Sex | 1.13 x10^9^ (0 – infinite) | .99 |
| PD-PRS / AD-PRS + Age + Sex, in poor vision group only (Low AD-PRS / Low PD-PRS) | | | |
|  | High AD-PRS / Low PD-PRS | 2.53 x10^9^ (0 – infinite) | .99 |
|  | Low AD-PRS / High PD-PRS | 3.56 x10^9^ (0 – infinite) | .99 |
|  | High AD-PRS / High PD-PRS | 5.64 x10^9^ (0 – infinite) | .99 |
|  | Age | 1.05 (0.95 – 1.16) | .80 |
|  | Sex | 0.85 (0.25 – 2.96) | .99 |

AD-PRS = Alzheimer’s Disease polygenic risk score, ApoE-E4 = Apolipoprotein E-E4 , GBA = glucosylceramidase beta, HR = hazard ratio, PD-PRS = Parkinson’s disease polygenic risk score, SE = standard error

**Supplementary Table 2. Models of Survival Time for progression to mild cognitive impairment in PD, using poor-vision as reference group**

| Model (comparator group) | Variable | HR (CI) | FDR corrected p-value |
| --- | --- | --- | --- |
| *GBA* mutations/Vision Group + Age + Sex (Poor vision / *GBA* negative) | | | |
|  | Good vision / *GBA* negative | 0.46 (0.16 – 1.35) | .25 |
|  | Good vision / *GBA* positive | 2.13 (0.56 – 8.07) | .39 |
|  | Poor vision / *GBA* positive | 1.27 (046 – 3.52) | .77 |
|  | Age | 1.06 (1.03 – 1.10) | **.002** |
|  | Sex | 0.88 (0.56 – 1.41) | .76 |
| APOE-E4 polymorphism/Vision Group + Age + Sex (Poor vision / ApoE-E4 negative) | | | |
|  | Good vision / APOE-E4 negative | 0.47 (0.25 – 0.90) | **.050** |
|  | Good vision / APOE-E4 positive | 0.83 (0.38 – 1.80) | .76 |
|  | Poor vision / APOE-E4 positive | 1.25 (0.71 – 2.20) | .60 |
|  | Age | 1.06 (1.02 – 1.10) | **.003** |
|  | Sex | 0.98 (0.61 – 1.58) | .99 |
| PD-PRS/Vision Group + Age + Sex (Poor vision / high PD-PRS) | | | |
|  | Good vision / low PD-PRS | 0.27 (0.12 – 0.58) | .**003** |
|  | Good vision / high PD-PRS | 0.71 (0.40 – 1.29) | .39 |
|  | Poor vision / low PD-PRS | 0.86 (0.50 – 1.47) | .74 |
|  | Age | 1.06 (1.03 – 1.10) | **.002** |
|  | Sex | 1.05 (0.64 – 1.70) | .94 |
| AD-PRS /Vision Group + Age + Sex (Poor vision /low AD-PRS) | | | |
|  | Good vision / low AD-PRS | 0.56 (0.26 – 1.20) | .24 |
|  | Good vision / high AD-PRS | 0.85 (0.46 – 1.58) | .76 |
|  | Poor vision / high AD-PRS | 1.91 (1.10 – 3.32) | **.050** |
|  | Age | 1.07 (1.03 – 1.10) | **.002** |
|  | Sex | 1.01 (0.62 – 1.63) | .99 |

**Supplementary Table 3. Estimates for sample sizes based on simulation for agent preventing milestone of conversion to PD-MCI or dementia**

| **Patient stratification method** | **Agent showing ~10% slowing in cognitive decline** | **Agent showing ~20% slowing in cognitive decline** | **Agent showing ~30% slowing in cognitive decline** |
| --- | --- | --- | --- |
| Unselected PD | n=4579 | n= 1255 | n= 530 |
| Unselected PD vision + PD PRS | n= 4220 | n= 1030 | n= 465 |
| Unselected PD vision + PD PRS + AD PRS | n= 3880 | n= 960 | n= 410 |
| PD poor vision unselected PRS | n= 3885 | n= 945 | n= 385 |
| PD poor vision  + AD PRS | n= 3180 | n= 715 | n= 325 |
| PD poor vision  + AD PRS + PD PRS | n= 3080 | n= 645 | n= 305 |

AD-PRS = Alzheimer’s Disease polygenic risk score, MoCA = Montreal Cognitive Assessment, PD = Parkinson’s disease, PD-PRS = Parkinson’s disease polygenic risk score

**Supplementary Figure 1**

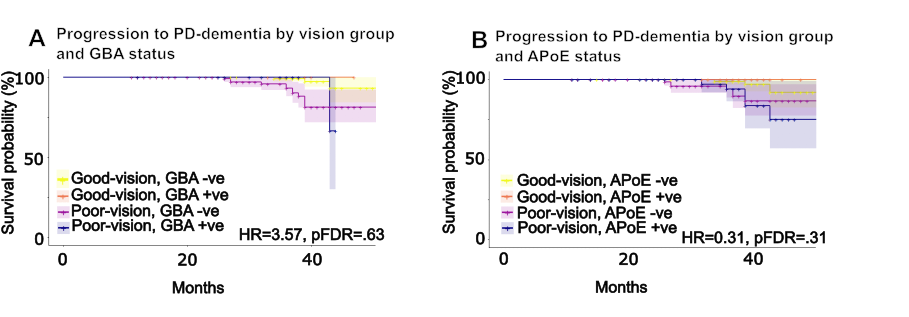

**Survival curves for progression to dementia in PD stratifying by visual performance and *GBA* and *APOE* high-risk mutations.**

Kaplan-Meier plots illustrating the probability of progression to PD dementia:

A) Divided by vision (good- versus poor-); and by *GBA* risk variant status (positive vs negative for the *GBA* risk variants *E365K* or *T408M*).

B) Divided by vision (good- versus poor-); and by *APOE* status (positive vs negative).

Dementia was defined using MoCA score <=21. Timeline is in months. Shaded area represents 95% CIs. Log rank score was calculated using Cox’s proportional hazards, with age-correction. Hazard ratios represent the difference between good vision/gene negative and poor vision/gene positive groups.

**Supplementary Figure 2**

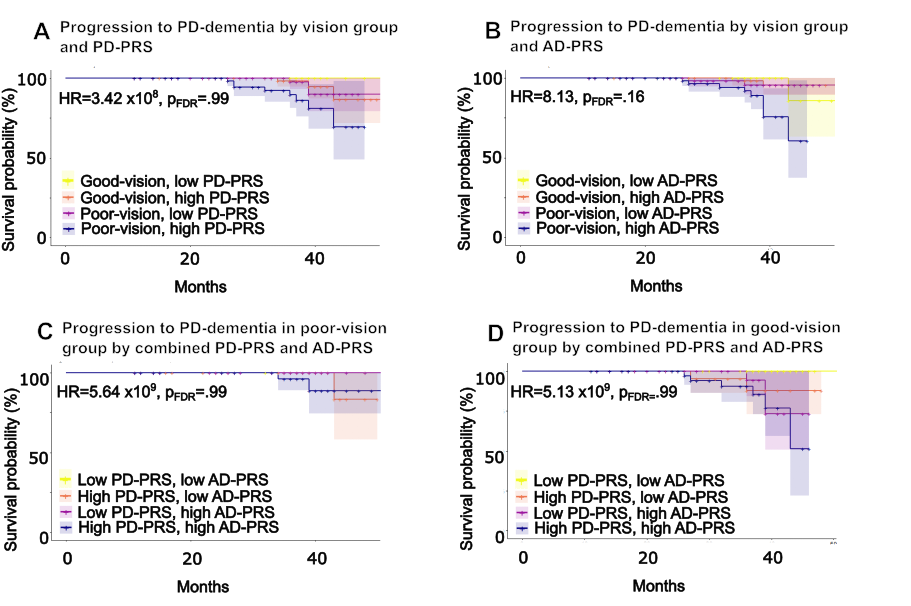

**Survival curves for progression to dementia in PD stratified by vision and polygenic risk scores.**

Kaplan-Meier plots illustrating the probability of progression to PD-dementia in:

A) PD patients, divided by vision (good- versus poor-); and by PD-PRS status (low vs high).

B) PD patients, divided by vision (good- versus poor-); and by AD-PRS status (low vs high).

C) PD good-vision patients, divided by combined PD-PRS and AD-PRS status (both low; high PD-PRS, high AD-PRS; both high).

C) PD poor-vision patients, divided by combined PD-PRS and AD-PRS status (both low; high PD-PRS, high AD-PRS; both high).

Dementia was defined using MoCA score <=21. Timeline is in months. Shaded area represents 95% CIs. Log rank score was calculated using Cox’s proportional hazards, with age-correction. Hazard ratios represent the difference between (A & B) good vision/low PRS and poor vision/high PRS groups or (C & D) low PD-PRS/low AD-PRS and high-PD-PRS/high AD-PRS.

AD, Alzheimer’s disease; PD, Parkinson’s disease; PRS, polygenic risk score.

**Supplementary Figure 3**

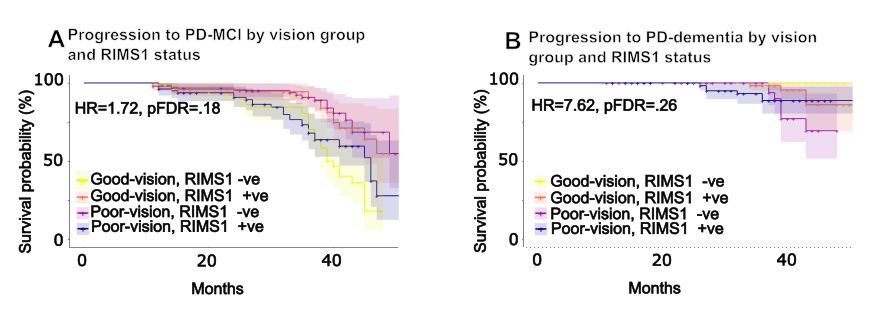

**Survival curves for progression to mild cognitive impairment and dementia in PD stratified by vision and *RIMS1* status.**

Kaplan-Meier plots illustrating the probability of:

A) Progression to mild cognitive impairment in PD patients divided by vision (good- versus poor-); and by *RIMS1* status (negative vs positive)
B) Progression to PD dementia in PD patients divided by vision (good- versus poor-); and by *RIMS1* status (negative vs positive)

MCI was defined using MoCA score <=26; Dementia was defined using MoCA score <=21. Timeline is in months. Shaded area represents 95% CIs. Log rank score was calculated using Cox’s proportional hazards, with age-correction. Hazard ratios represent the difference between good vision/*RIMS1* negative and poor vision/*RIMS1*positive.

PD-MCI = Mild cognitive impairment; MoCA = Montreal Cognitive Assessment, PD = Parkinson’s disease.

**Supplementary Figure 4**

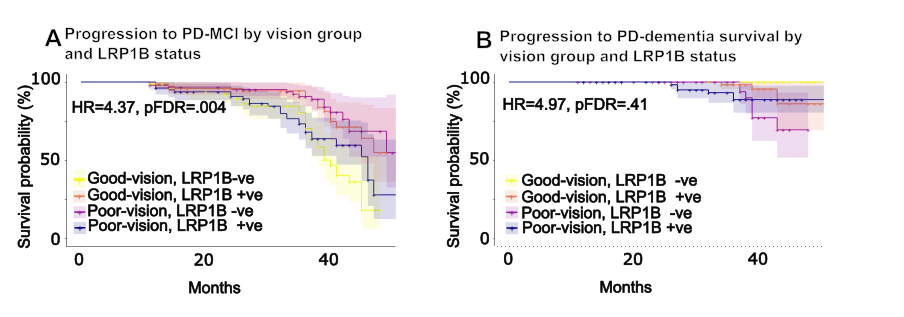

**Survival curves for progression to mild cognitive impairment and dementia in PD stratified by vision and *LRP1B* status.**

Kaplan-Meier plots illustrating the probability of:

A) Progression to mild cognitive impairment in PD patients divided by vision (good- versus poor-); and by *LRP1B* status (negative vs positive)
B) Progression to PD dementia in PD patients divided by vision (good- versus poor-); and by *LRP1B* status (negative vs positive)

MCI was defined using MoCA score <=26; Dementia was defined using MoCA score <=21. Timeline is in months. Shaded area represents 95% CIs. Log rank score was calculated using Cox’s proportional hazards, with age-correction. Hazard ratios represent the difference between good vision/LRP1B negative and poor vision/LRP1B positive.

MCI = Mild cognitive impairment; MoCA = Montreal Cognitive Assessment, PD = Parkinson’s disease.

**References**

1.	Weil RS, Schwarzkopf DS, Bahrami B, et al. Assessing cognitive dysfunction in Parkinson's disease: an online tool to detect visuo‐perceptual deficits. *Movement Disorders.* 2018;33(4):544-553.

2.	Jiang Z, Wang L, Li C, Xia J, Jia H. A practical simulation method to calculate sample size of group sequential trials for time-to-event data under exponential and Weibull distribution. 2012.
